# Multivariate Classification of First-Episode Schizophrenia Spectrum Psychosis using EEG Microstate Dynamics

**DOI:** 10.64898/2026.02.18.26346582

**Authors:** Aron T. Hill, Neil W. Bailey, Talitha C. Ford, Jarrad A.G. Lum

## Abstract

**Background:** EEG microstates provide a window into rapid, large-scale brain network dynamics. Despite showing alterations in schizophrenia, evidence in first-episode schizophrenia spectrum psychosis (FESSP) is limited. We assessed whether microstate temporal and transition features could identify a multivariate signature of FESSP, and whether these dynamics can track symptom severity.

**Methods:** Resting-state EEG was analysed in 69 participants (FESSP n=41, mean age: 22.49 years; healthy controls n=28, mean age: 21.33 years). Twenty-eight microstate temporal and transition features were extracted across microstate classes (A-D). Group classification accuracy was assessed using a linear support vector machine with stratified cross-validation and permutation testing. Within the FESSP group, we further assessed associations between microstate features and clinical scores using the Brief Psychiatric Rating Scale (BPRS), Scale for the Assessment of Positive Symptoms (SAPS), and Scale for the Assessment of Negative Symptoms (SANS).

**Results:** Multivariate microstate features provided above-chance discrimination of FESSP from controls (balanced accuracy=0.644; AUC=0.688; p=0.030). However, when comparing individual features between groups, no feature survived multiple-comparison correction consistent with characterisation of FESSP via a distributed multivariate pattern across correlated features. Within the FESSP group, microstate dynamics were most strongly linked to negative symptoms, with higher SANS scores associated with shorter microstate D durations (ρ=-0.507, pFDR=0.020) and higher occurrence of microstates A and B (ρ=0.434–0.443, pFDR=0.042). BPRS-18 and SAPS showed no associations with any features.

**Conclusions:** Using EEG microstate temporal and transition features with multivariate classification, we identified a pattern that differentiated FESSP from controls and showed selective associations with negative symptom severity.

## INTRODUCTION

Psychotic disorders, including schizophrenia spectrum disorders, are a leading cause of long-term disability and show increasing global prevalence (1,2). Psychosis is characterised by impaired reality testing, including hallucinations and delusions, which can be highly functionally disruptive (3). As outcomes are strongly shaped by processes occurring early during the illness course, early detection and intervention are critical (4-6). This motivates the need for objective, scalable markers in first-episode schizophrenia spectrum psychosis (FESSP) that capture early brain dysfunction and link them to clinical symptoms (7,8). Electroencephalography (EEG) is well suited to this goal as a non-invasive, cost-effective, and scalable method (9-11), that provides millisecond resolution of brain dynamics.

Accumulating evidence implicates disrupted large-scale network dynamics as contributing to the clinical symptoms in psychosis and schizophrenia spectrum disorders, including FESSP (12-15). EEG-derived microstates offer one approach to quantify these dynamics. Microstates provide a time-resolved window into rapid switching between the activation of large-scale functional brain networks, complimenting conventional spectral and connectivity analyses by capturing the temporal organisation of global electrical field patterns (16-18). They segment the continuous EEG signal into brief epochs of quasi-stable scalp potential topographies, which typically last between tens to hundreds of milliseconds (16,19).

A small set of canonical microstate classes (most commonly A-D) have been reliably observed across both neurotypical and clinical cohorts. These classes reflect recurring quasi-stable scalp potential topographies that typically persist for tens of milliseconds before rapidly transitioning to another class (16,20,21). Each microstate class is thought to reflect transient coordinated configuration of global network activity, with the rapid alterations between microstate classes indicative of large-scale changes in functional brain organisation (16,22). Microstates are separated by their topographical patterns, with maps of inverted polarities typically clustered together, as they are assumed to arise from the same underlying neural generators. Microstate A is typically characterised by a left-posterior-negative-to-right-frontal-positive (or vice versa) topographical orientation, microstate B shows the mirrored pattern to Microstate A, with an opposite right-to-left orientation, microstate C shows a predominantly anterior-positive-posterior-negative (or vice versa) configuration, and microstate D shows a frontocentral predominance (see Figure 1A) (16).

**Figure 1:**
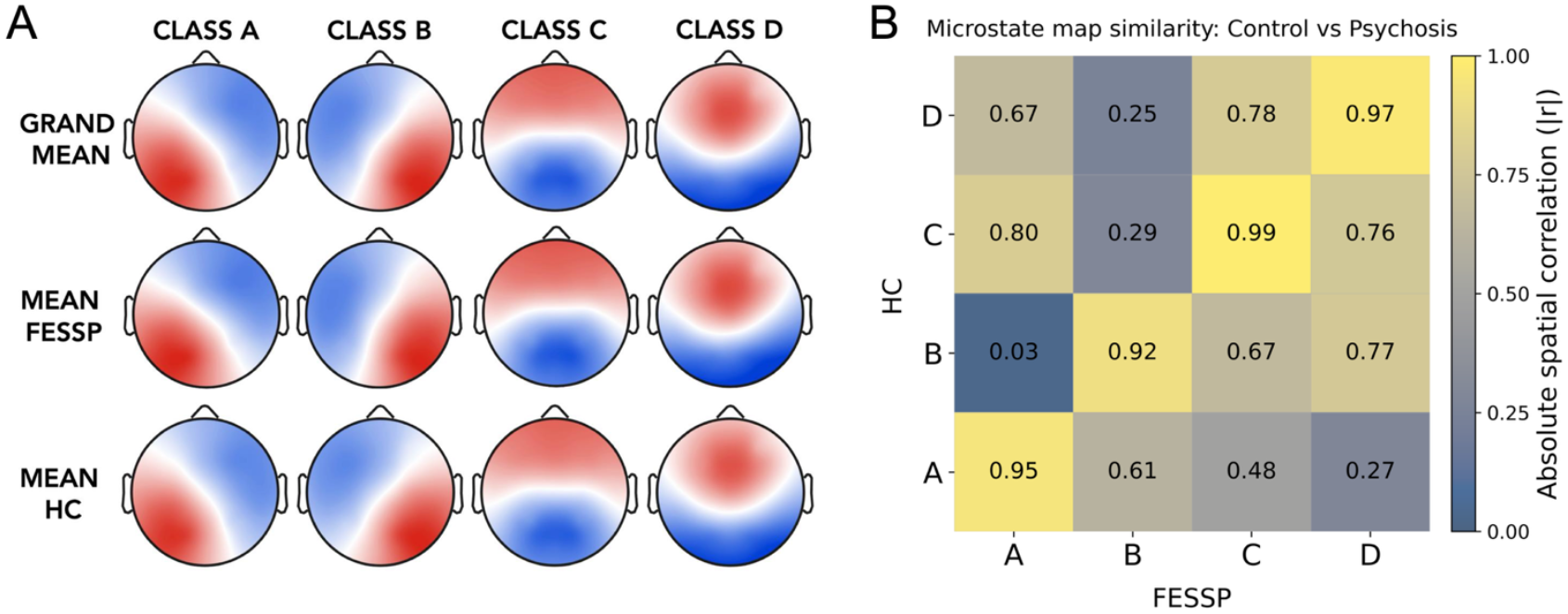
A) Topographic plots of the four EEG microstate classes (A-D) for the Grand Mean (top), first-episode schizophrenia spectrum psychosis (FESSP) group (middle), and healthy control (HC) group (bottom). The topographies closely resembled the orientations previously reported in the literature: Microstate A: right anterior to left posterior; Microstate B: left anterior to right posterior; Microstate C: frontal to occipital; Microstate D: medial anterior to occipital. Note, microstates were derived using a polarity invariant (modified K-means) clustering algorithm. B) Spatial correlations between the FESSP and HC microstate topographies.

Specific temporal parameters such as average *duration, occurrence* rate, time *coverage*, and *explained variance*, as well as sequence characteristics such as transition probabilities, can then be calculated within each microstate class. Average duration indexes how long a microstate remains stable, while occurrence rate indexes how frequently a microstate appears. Time coverage indicates the amount of time the microstate is present across the total time of each EEG recording, and finally, explained variance reflects the percentage of total variance in the data explained by a given microstate (a measure driven by both the amplitude and time coverage of each microstate, relative to other microstates) (23,24). In addition, transition probabilities quantify the likelihood of switching from one specific microstate class to another class at the next time step, thus capturing the preferred sequencing of states, a measure that complements duration, occurrence, coverage, and explained variance by describing how specific microstate classes follow one another rather than simply how long or how often they occur (25,26).

Across the schizophrenia spectrum, microstate abnormalities have been most frequently reported for classes C and D (27-29). A comprehensive meta-analysis found increased occurrence of microstate C and shortened duration of microstate D (28). Evidence from patients and unaffected siblings further supports microstates C and D as candidate endophenotypes (29). For example, in a cohort including patients with schizophrenia, their unaffected siblings, healthy controls, and a first-episode psychosis group, both patients and siblings showed increases across several temporal features for microstate C and reductions for microstate D relative to controls (29). However, microstate temporal parameters did not differ between first-episode psychosis and matched chronic patients (29). While microstate alterations in psychosis are most consistently reported for classes C and D (30), findings for classes A and B are more heterogeneous, with meta-analyses suggesting no consistent effect for class A and only weak evidence for class B duration, and several early-phase studies indicating that A and B dynamics may vary with clinical stage and antipsychotic exposure (28).

Importantly, even relatively consistent group-average effects do not necessarily yield useful individual-level signatures, thus limiting potential diagnostic value. This motivates multivariate approaches that test the possibility of individual classification, as well as generalisation under cross validation (31,32). Machine learning can bridge this gap by using distributed, multivariate patterns that may be subtle or unreliable at the single-feature level but become predictive when combined across features (32,33). In psychiatric neuroscience, supervised classifiers are increasingly used to evaluate whether candidate neural markers support reliable individual-level discrimination, but the translational value of such models depends critically on rigorous out-of-sample evaluation and the avoidance of optimistic bias from analytic flexibility (34).

In the present study, we tested whether commonly reported EEG microstate temporal and transition features can distinguish individuals with FESSP from healthy controls using a cross-validated linear support vector machine (SVM). To link discrimination-relevant microstate dynamics with clinical phenotype, we also examined associations between microstate features and symptom severity scales in the FESSP group. This combined decoding and brain-behaviour framework builds on emerging evidence that microstate dynamics support multivariate discrimination in schizophrenia, while explicitly addressing common evaluation pitfalls emphasised in the broader decoding literature (35). We hypothesised that microstate dynamics would reliably discriminate FESSP from healthy controls under stratified cross-validation, and that clinical symptoms would be associated with more atypical microstate stability and sequencing.

## METHODS

### Participants

This study analysed a pre-collected dataset from OpenNeuro (Dataset: ds003944). For a detailed overview of the experimental methodology see Phalen et al (36). The full dataset comprised 82 participant files (age-range: 12.8-35.9 years). After excluding duplicate records and participants with missing or incomplete EEG or clinical/demographic data, or excessively poor EEG quality, 69 participants remained (41 FESSP, 28 healthy control [HC]). FESSP diagnoses were established using DSM-IV criteria. Group demographics and medication details are presented in Table 1. All participants provided informed consent and ethical approval for the original data collection was provided by the University of Pittsburgh’s International Review Board. Deakin University granted an ethics exemption for secondary analysis.

**Table 1:** Participant Demographics.

|  | Healthy control | FESSP |
| --- | --- | --- |
| **N (Male: Female)** | 28 (16:12) | 41 (28:13) |
| **Age, mean (SD)** | 21.33 (3.95) | 22.49 (5.28) |
| **Antipsychotic medicated, yes/no/unknown (n)** | - | 25/14/2 |
| **BPRS-18, mean (SD)** | - | 44.37 (8.07) |
| **SAPS, mean (SD)** | - | 22.54 (12.93) |
| **SANS, mean (SD)** | - | 30.80 (15.97) |

*Note:* FESSP: first-episode schizophrenia spectrum psychosis, SD; standard deviation, BPRS: Brief Psychiatric Rating Scale, SAPS: Scale for the Assessment of Positive Symptoms, SANS: Scale for the Assessment of Negative Symptoms.

### Clinical Rating Scales

As part of the original study, FESSP participants completed several clinical assessments (see Phalen et al (36)). Here, we extracted scores from three widely used measures: the 18-item Brief Psychiatric Rating Scale (BPRS) (37), the Scale for the Assessment of Positive Symptoms (SAPS) (38), and the Scale for the Assessment of Negative Symptoms (SANS) (39). The BPRS indexes overall psychopathology spanning positive, negative, affective, and activation symptoms, with higher total scores indicating greater symptom severity (40). The SAPS quantifies positive symptom severity across hallucinations, delusions, bizarre behaviour, and formal thought disorder; while the SANS quantifies negative symptom severity across affective blunting, alogia, avolition or apathy, anhedonia or asociality, and attention, with higher scores indicating greater severity in each domain (41).

### EEG Data Acquisition

The EEG dataset consisted of five minutes of eyes-open resting-state recordings acquired with a Neuromag Vectorview system (Elekta, Helsinki, Finland) at 1 kHz. Participants sat upright in a magnetically shielded room and fixated on a central cross throughout the recording (36). Data were acquired with a low-impedance 60-channel cap. Magnetoencephalography (MEG) data were acquired in the same session but are not presented here (reported in Phalen et al (36)).

### EEG Pre-Processing

EEG data were pre-processed in MATLAB (v9.10.0) using the RELAX (Reduction of Electroencephalographic Artifacts; v1.1.5) software (42,43). As part of this pipeline, data were bandpass filtered (0.5-80 Hz; fourth-order zero-phase Butterworth filter) with a notch filter (57-63 Hz) to attenuate line noise. Bad channels were removed using a multi-step process incorporating the ‘findNoisyChannels’ function from the PREP pipeline (and later re-interpolated using spherical interpolation; mean removed: FESSP=6.41 [SD=3.45], HC=6.46 [SD=4.27]) (44). Multi-channel Wiener filters were then used to reduce blinks, muscle activity and eye movements (45), followed by robust average re-referencing and wavelet-enhanced independent component analysis (46), with artifact components identified using ICLabel (47). Cleaned continuous data were segmented into non-overlapping four-second epochs. Because microstate estimation is sensitive to residual artifacts (48), this validated pipeline was used to minimise any artifacts prior to analysis. Finally, in preparation for the microstate analysis, the data were further bandpass filtered (1-40 Hz), and down-sampled to 250 Hz to reduce computational burden (23,49).

### Microstate Analysis

Microstate analysis was conducted in MATLAB using MICROSTATELAB (26) with identical settings for FESSP and HC groups. Individual microstate maps were identified using the modified k-means clustering algorithm (20 random restarts) applied to maps of the peak global field power (GFP) to maximise signal-to-noise ratio (50). Prior to clustering, maps were normalised, and polarity was ignored (opposite polarity treated as equivalent) (26). Because individual clustering yields maps in an arbitrary order, MICROSTATELAB’s hierarchical averaging and sorting procedures were used to derive group-and sample-level template maps. To permit comparison with canonical microstates, grand-average maps were automatically sorted with respect to the published template maps of Koenig et al. (50) and microstate classes in the 4-class solution were labelled A, B, C, and D according to their topographic similarity to these templates. Analyses were restricted to the four-class solution to align with the canonical resting-state microstate literature in which the four canonical classes A-D are the most robust and consistently replicated, thus maintaining generalisability (16,50,51). This approach also limits the number of derived features and associated number of multiple comparisons for the statistical analyses, which is important given the sample size. Although additional microstate classes have been reported, the four canonical microstates are robust and widely recognised across both healthy and clinical populations (16,20).

Temporal microstate parameters were extracted by backfitting the subject-specific microstate templates to each participant’s EEG using MICROSTATELAB (26). Backfitting was restricted to GFP peaks, with class labels extended to intervening samples using the nearest neighbour criterion.

Although MICROSTATELAB outputs many measures; our analyses focussed on a subset of commonly reported features, namely average duration, occurrence rate, time coverage, and explained variance for each separate microstate (a total of 16 features), plus the pairwise transition probabilities between each of the microstate classes (12 features, making a total of28 features).

### Statistical Analysis

#### Multivariate Classification

To test whether the joint microstate feature pattern discriminated the FESSP from HC groups at the single-subject level, we trained a linear support vector machine (SVM) on the 28-dimensional microstate feature space with group encoded as a binary label (FESSP=1, HC=0). Analyses were implemented in Python using SciPy (52), Statsmodels (53), and scikit-learn (54). Features were standardised (zero mean, unit variance) via StandardScaler within a scikit-learn pipeline, ensuring scaling parameters were estimated within the training fold only. The classifier used a linear kernel with balanced class weights to account for unequal sample size between groups (55).

#### Cross-Validation, Performance Metrics and Permutation Testing

Classification accuracy performance was estimated using a stratified 5-fold cross validation with a fixed random seed, preserving the FESSP:HC ratio within each fold. Five folds were selected as a pragmatic compromise between bias and variance for the present study’s sample size, providing a commonly used cross-validated estimate of performance on held-out data and typically lower variance than leave-one-out cross validation approaches (56,57). For each fold, we computed balanced accuracy and receiver operating characteristic (ROC) area under the curve (AUC).

Statistical significance was assessed using a permutation test (1,000 iterations) to obtain a null distribution of balanced accuracy scores. The group labels were randomly permuted and for each permutation the entire stratified 5-fold cross validation pipeline was repeated and balanced accuracy recomputed. The permutation *p*-value was defined as the proportion of permuted balanced accuracies that were at least as large as the observed balanced accuracy on the true labels.

#### Univariate Feature Characterisation and Model Interpretation

Following classification, we performed descriptive univariate analyses for each microstate feature to contextualise multivariate performance. Group differences were tested using Welch’s *t*-tests and quantified with Hedges’ g, with false discovery rate (FDR) correction applied across the 28 features. We also computed univariate ROC AUC for each feature as a descriptive index of single-feature discriminability. To aid interpretation of the multivariate classifier, we also computed out-of-fold SHapley Additive exPlanations (SHAP) values for the linear SVM decision function, which decompose each held-out prediction into additive per-feature contributions relative to the model’s expected value (58). Global feature importance was summarised using the mean absolute SHAP value across participants.

#### Associations with Clinical Scores

As an additional exploratory analysis, we assessed for any associations between the various microstate temporal and transition features and clinical symptom severity in the FESSP group. Using the same feature set as the classification analysis, we computed Spearman correlations between each microstate feature and scores on the BPRS, SAPS, and SANS. This approach complements the multivariate decoding by testing whether microstate parameters vary systematically with clinical phenotype, rather than simply differentiating patients from controls. To control for multiple testing, *p*-values were FDR-corrected (Benjamini-Hochberg) (59) across the 28 feature-wise correlations within each clinical scale. Finally, we also computed Pearson’s spatial correlations for each of the four microstate classes between the FESSP and HC recordings to assess for topographic similarity between the two groups.

## RESULTS

Mean microstate maps for the FESSP and HC groups separately, as well as the Grand Mean (mean across both groups) are shown in in Figure 1. Maps closely resembled canonical maps from the literature (16,20,24,60). The four microstate classes accounted for 70.46% (HC) and 71.08% (FESSP) of the total global variance, broadly consistent with previous reports (16,61). Figure 2 provides an overview of the temporal parameters for each microstate separately for the FESSP and HC groups.

**Figure 2.**
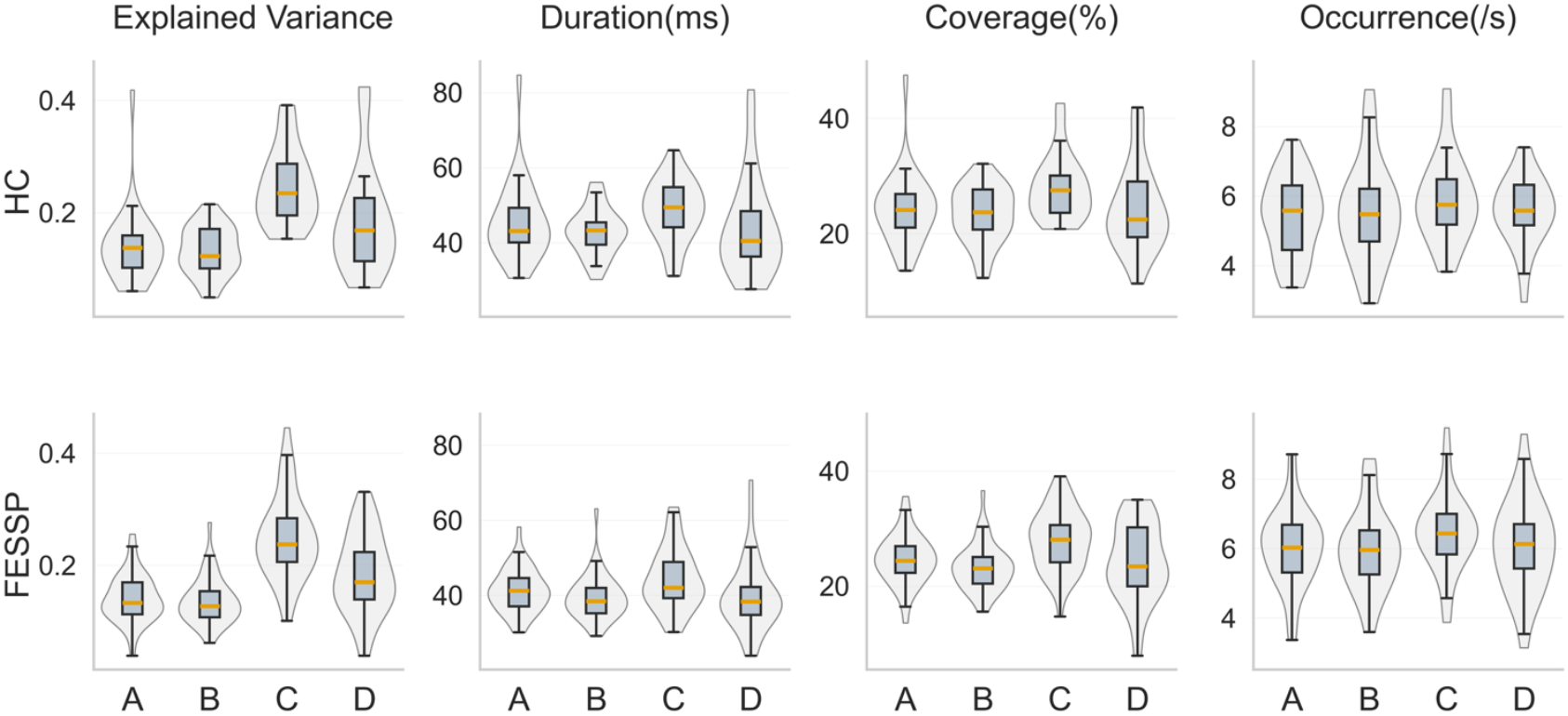
Distribution of microstate features by group (FESSP, HC) and microstate class (A-D). Violin plots show the distribution of individual-level microstate measures for each class separately for healthy controls (HC) and first-episode schizophrenia spectrum psychosis (FESSP). Overlaid boxplots indicate the median and interquartile range, with whiskers showing the range excluding outliers. Panels depict class-wise individual explained variance (IndExpVar; labelled GEV), mean microstate duration (ms), coverage (% of time spent in each microstate), and mean occurrence rate (events/s).

### Multivariate classification of psychosis status

The SVM indicated that the microstate features provided above-chance discrimination of FESSP from HCs. The classifier achieved a balanced accuracy of 0.644 (95% CI=0.513-0.788) and ROC AUC of 0.688 (95% CI=0.497-0.831; Figure 3B). In permutation testing, the observed balanced accuracy exceeded the null distribution (*p*=0.030; Figure 3A), indicating that the observed classification performance was unlikely under the null hypothesis of no association between multivariate microstate dynamics and group membership.

**Figure 3:**
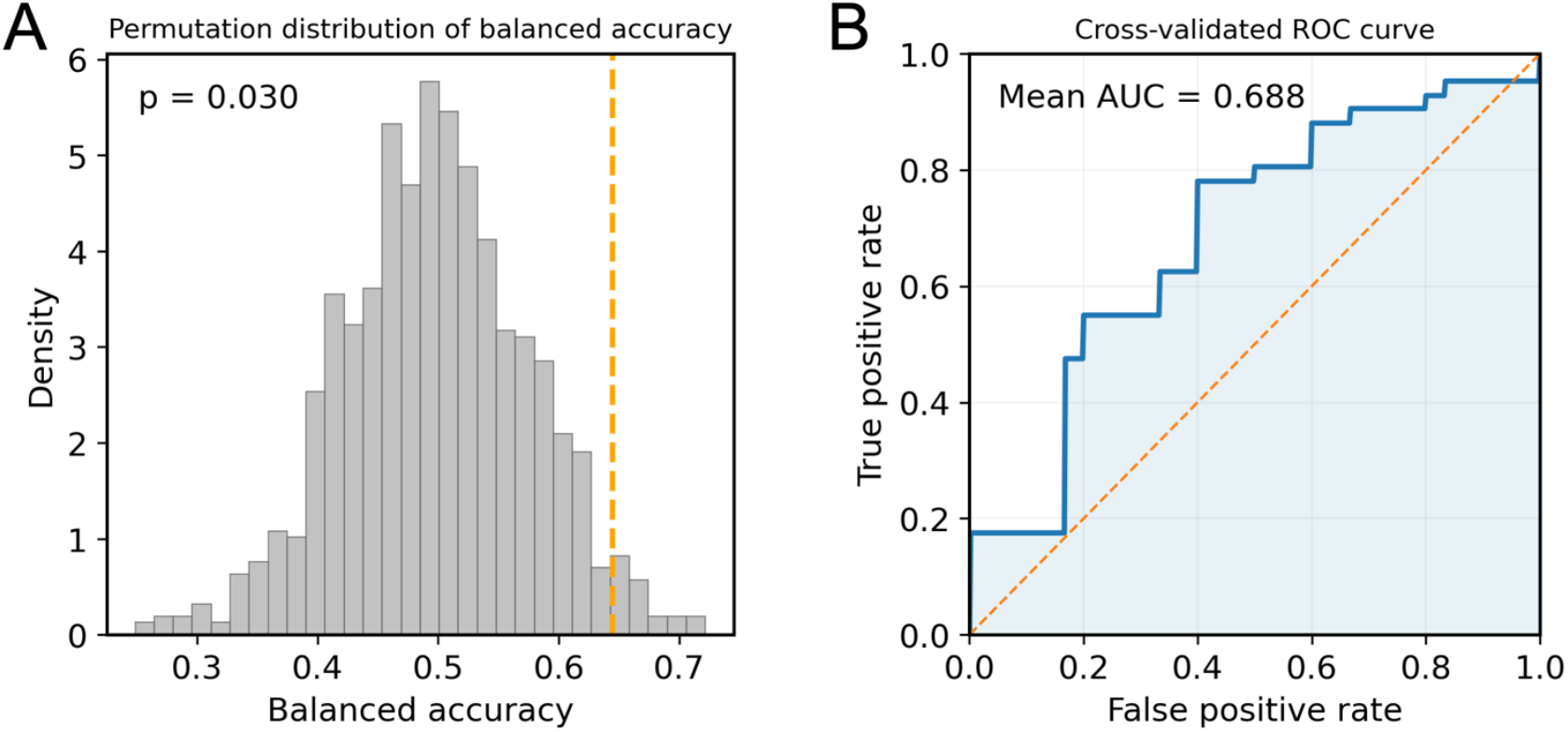
Support vector machine (SVM) classification performance and univariate feature discrimination. A) Histogram shows the distribution of balanced accuracies obtained from 1,000 label permutations, with the full stratified 5-fold cross-validation pipeline repeated for each shuffle. The dashed vertical orange line marks the observed balanced accuracy for the true healthy controls (HC) versus first-episode schizophrenia spectrum psychosis (FESSP) labels. The permutation *p*-value is the proportion of permuted accuracies at least as large as the observed value. B) Cross-validated receiver operating characteristic (ROC) curve for the linear SVM. The solid blue line shows the mean ROC curve across the 5 test folds, computed from fold-wise SVM decision values (decision_function) and averaged after interpolation onto a common false positive rate grid. The dashed diagonal orange line indicates chance performance (AUC=0.5). The mean fold-wise area under the ROC curve (AUC) is reported on the panel.

Univariate analyses showed nominal effects that were most evident for temporal parameters, with shorter mean durations for classes A-C in FESSP (FESSP <HC; Hedges’ g =∼0.53-0.56) and higher mean occurrences for classes A, C, and D (FESSP >HC; Hedges’ g =∼0.42-0.55; full results in Supplemental Table S1). However, no individual feature survived FDR correction (all pFDR≥0.288) consistent with the difference between FESSP characterised by a distributed, multivariate signal rather than a single dominant feature. Single-feature discriminative performance was similarly modest, with univariate ROC AUCs summarised in Figure 4A. Finally, mean absolute SHAP values, which index the magnitude of each feature’s contribution, indicated a distributed set of influential predictors spanning explained variance (IndExpVar_B/C, where more positive values increased the likelihood that the model would classify a participant as FESSP), temporal parameters (MeanOccurrence_B; MeanDuration_A/C, where more negative values increased the likelihood that the model would classify a participant as FESSP; Coverage_D), and transition probabilities (multiple OrgTM transitions, where lower values for transitions from C>B, A>D, and B>A associated with increased likelihood that the model would classify a participant as FESSP, and vice versa for transitions from D>A, A>B, A>C, C>D, and C>A). The wide range of influential predictors is consistent with classification being supported by a multi-feature pattern, rather than a single dominant marker (Figure 4B). An additional detailed ‘beeswarm’ plot of SHAP values is provided in the Supplemental Materials (Figure S1).

**Figure 4:**
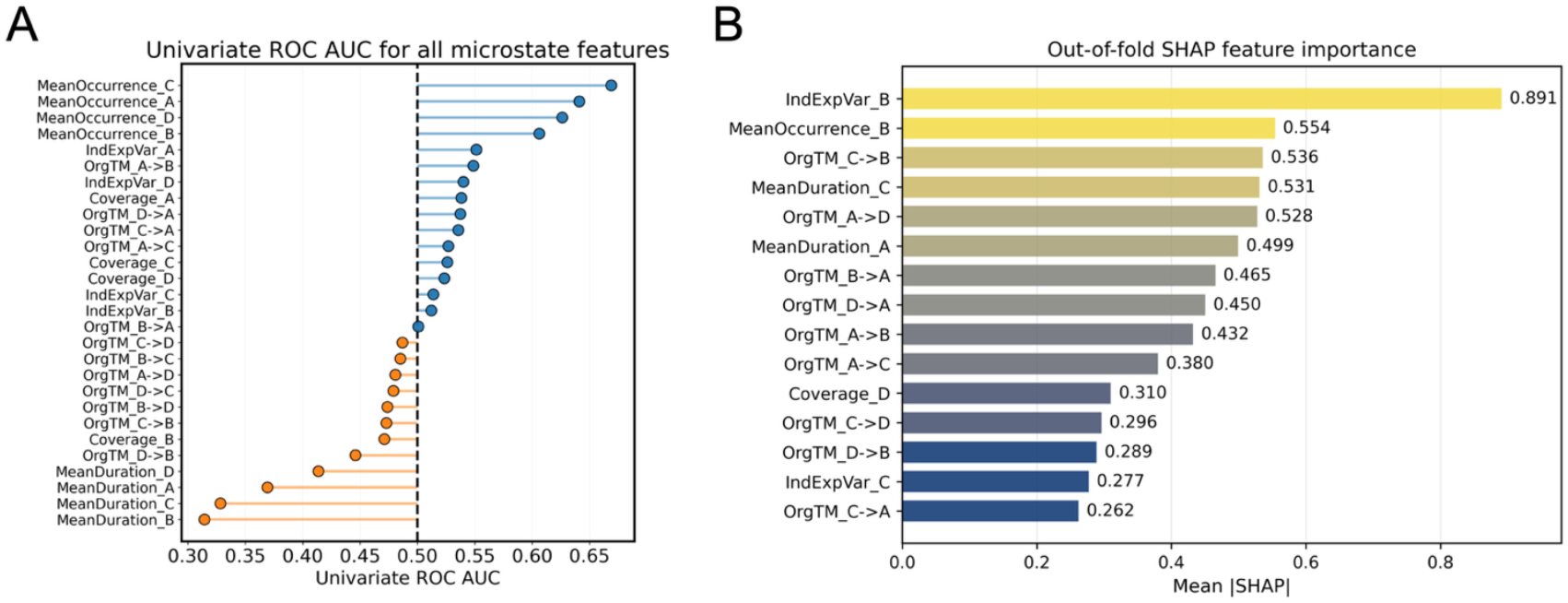
A) Summary of univariate receiver operating characteristic (ROC) area under curve (AUCs) for all microstate features examined using a lollipop plot. Each lollipop represents the ROC AUC of a single feature computed across the full sample for discriminating healthy controls (HC) versus first-episode schizophrenia spectrum psychosis (FESSP). Features are ordered by AUC. The dashed vertical line at AUC=0.5 indicates chance level discrimination. Blue features have AUC≥0.5, indicating higher values in FESSP compared to HC under the chosen coding of the positive class, whereas orange features have AUC<0.5, indicating discrimination in the opposite direction.

Importantly, AUC values below 0.5 reflect reversed directionality (not weaker discriminative information) and can be converted to an equivalent discriminability as 1−AUC. Note, this figure is designed for descriptive interpretation only and does not reflect cross-validated performance or multiple comparison control. B) Out-of-fold SHAP feature importance for the multivariate linear SVM classifier. SHAP values were computed within each cross-validation fold using models fit on the training set and evaluated on the held-out fold, yielding additive per-feature contributions to the SVM decision function for each participant. Bars show mean absolute SHAP values across participants (top 15 features), indexing the magnitude of each feature’s contribution to the multivariate function. Larger values indicate greater influence on the classifier’s predictions (but do not index the direction of the association. Additional beeswarm plots, which provide further information on directionality, are provided in the Supplemental Materials [Figure S1]).

### Associations with clinical ratings

Within the FESSP group, no microstate feature showed an association with overall psychopathology (BPRS; all pFDR ≥0.215) or positive symptoms (SAPS; all pFDR≥0.712) after FDR correction across the 28 features (Supplemental Tables S3-4). In contrast, negative symptom severity (SANS) was robustly associated with specific microstate dynamics. Higher SANS scores were associated with shorter mean duration of microstate D (ρ=-0.507, p=0.001, pFDR =0.020) and greater occurrence of microstates A (ρ=0.434, p=0.005, pFDR =0.042), and B (ρ=0.443, p=0.004, pFDR =0.042; Figure 5). Several other associations with SANS scores showed consistent directionality but did not survive correction (pFDR=∼0.054-0.12), including reduced C-to-D transitions (ρ=-0.41, p=0.008, pFDR=0.054) and increased A-to-B transitions (ρ=0.399, p=0.01, pFDR=0.055; for all correlations see Supplemental Table S5 and Figure S2). Collectively, these results suggest that microstate dynamics were more strongly linked to negative symptom expression than to positive symptoms or overall symptom burden.

**Figure 5.**
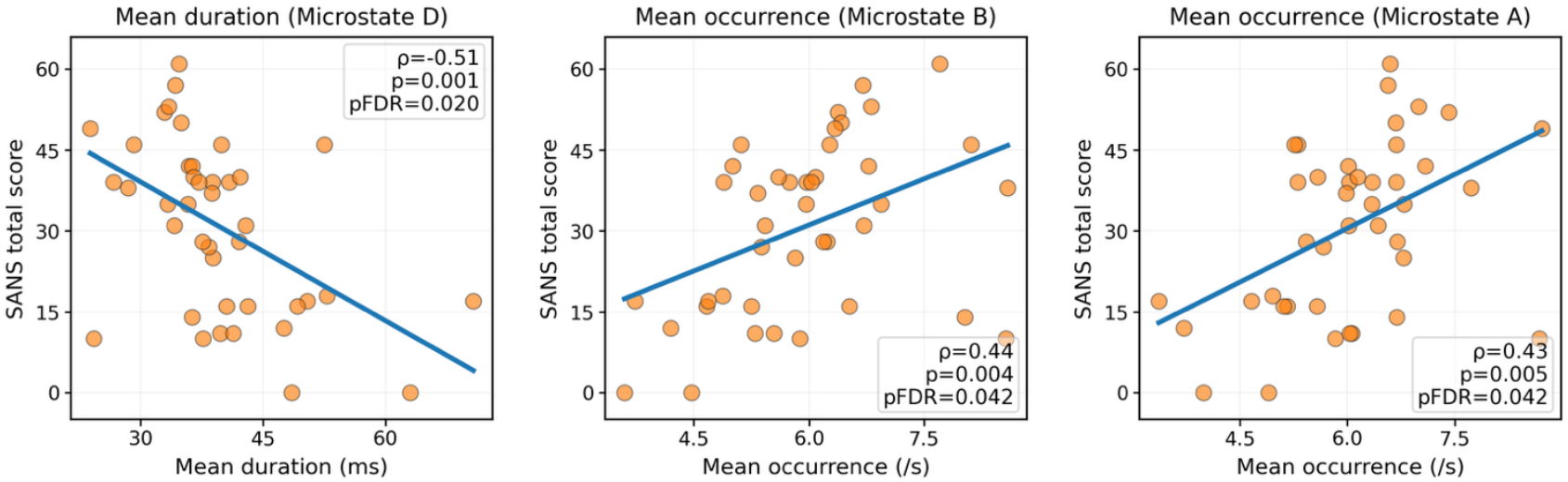
Associations between microstate features and negative symptom severity. Scatter plots show relationships between selected EEG microstate features and Scale for the Assessment of Negative Symptoms (SANS) total scores in the first-episode schizophrenia spectrum psychosis (FESSP) group. Each panel displays individual participant values and the least-squares regression line (blue). Panels are annotated with Spearman correlation coefficients (ρ), nominal *p-*values, and FDR-corrected *p*-values (pFDR). Mean occurrence for microstates A and B showed a positive moderate association with SANS scores, while mean duration for microstate D showed a moderate-to-strong negative association.

## DISCUSSION

We found that a compact set of neural activity patterns represented by microstate temporal and transition features extracted from resting-state EEG can classify FESSP compared to HCs with above chance balanced accuracy. The linear SVM, trained on 28 features across microstates A-D, achieved modest but statistically significant out-of-sample discrimination (balanced accuracy=0.644; AUC=0.688; p=0.030). Notably, no individual microstate parameter survived multiple-comparison correction in univariate analyses, indicating that discrimination was driven by a distributed multivariate pattern across correlated features rather than a single dominant effect. Consistent with this, the out-of-fold SHAP values indicated that classification was supported by a distributed set of contributors spanning explained variance, temporal parameters, and transition probabilities, rather than any single dominant feature. In addition, our exploratory analyses of clinical symptom associations suggests a relationship between negative symptom severity and microstate features, with higher SANS scores associated with shorter microstate D duration and increased occurrence of microstates A and B (surviving FDR-correction across all 28 comparisons). By contrast, SAPS showed no robust associations, and BPRS effects were limited to nominal, non-FDR-significant correlations.

Our findings broadly align with the wider schizophrenia microstate literature, which most consistently reports alterations to microstates C and D, with fewer and more heterogeneous findings for classes A and B (20). For example, a recent meta-analysis found that, compared to controls, individuals with psychosis showed no difference in microstates A or B, with the exception of the duration of microstate B in FESSP (20), whereas increases in occurrence and coverage in microstate C have been repeatedly reported across schizophrenia meta-analyses (20,28,29).

Importantly, this work extends the psychosis microstate literature in two ways. First, we evaluated whether microstate features support single-subject discrimination of diagnostic group membership under cross-validation testing, rather than relying solely on group-average differences that may not translate to individual classification (32,57). Specifically, we used a regularised linear SVM, which is well suited to modest sample sizes and correlated feature sets, and evaluated performance with stratified 5-fold cross-validation. Second, we limited analytic degrees of freedom by pre-specifying a compact, literature-motivated feature set and a single classifier with fixed hyperparameters (so as to reduce the risk of optimistic performance inflation from iterative model tweaking) (62,63), and by evaluating statistical significance with label permutation testing in which the entire cross-validation pipeline was repeated for each shuffle. This design aligns with recommended practice for predictive modelling in neuroimaging and electrophysiology, helping mitigate optimistic bias from analytic flexibility and providing a more defensible estimate of out-of-sample performance (34,35).

### Associations with Clinical Symptom Severity

While we assessed associations between microstate features and three measures of psychiatric symptoms (BPRS, SAPS, SANS), the strongest brain-symptom relationships emerged for negative symptoms (SANS scores). After FDR correction, higher SANS scores were significantly associated with shorter microstate D duration and higher occurrence of microstate A and B. The association between higher SANS scores and shorter microstate D duration is consistent with evidence that microstate D shows reduced temporal in schizophrenia (28). Microstate D has been linked to fronto-parietal BOLD-fMRI activity (17), a network implicated in attentional control, suggesting that reduced sustained engagement of top-down control states in FESSP (shorter microstate D duration) may relate to negative symptoms. This aligns with conceptualisations of negative symptoms (e.g., anhedonia, reduced goal-directed behaviour, diminished affect and social drive) (64) as impaired capacity to maintain attentional control in pursuit of internally represented goals (65,66). This finding is also consistent with evidence that microstate D’s temporal stability is reduced in schizophrenia spectrum disorders (25,28,67-69) and may reflect altered engagement of attention or cognitive control related network states.

In parallel, higher SANS scores were associated with higher occurrence of microstates A and B, commonly linked to auditory and visual resting-state networks, respectively (17). Previous studies have reported increased microstate A occurrence in schizophrenia, including FESSP (25,68,70), with de Bock et al. proposing that increases in microstate A parameters might represent a non-specific marker of general pathology (70). However, findings for microstate A in FESSP are mixed, and reductions in microstate A occurrence have also been reported relative to controls (30). Given we only found significant brain-behaviour relationships for negative symptoms (SANS) but not for BPRS or SAPS, one interpretation is that increased occurrence of microstates A and B may be more closely related to negative symptom burden in this dataset. Mechanistically, this pattern is consistent with a redistribution of microstate dynamics away from temporally stable control or attention-related states and towards more frequent engagement of sensory network states, aligning with models of psychosis as a disorder of dysconnectivity and impaired top-down coordination of distributed brain systems (71-73).

### Clinical Significance

Although SVM classification performance was relatively modest, the above chance cross-validation classification accuracy indicates that resting-state microstate dynamics contain information that can be used to differentiate FESSP from HCs. Replication in larger samples is needed, but these features could potentially contribute to multi-modal risk stratification approaches, with EEG offering an accessible and low-cost method for obtaining clinically relevant information on early psychosis neurobiology. Larger studies could also explore the multivariate interactions that drive classification accuracy, with the hope of revealing novel mechanistic markers of differences in FESSP.

The association between microstate dynamics and negative symptom severity is of particular clinical importance because negative symptoms are a major driver of long-term functional disability in schizophrenia, and are often less responsive to treatment (64,74). This suggests that microstate markers, particularly reduced temporal stability of microstate D and increase occurrence of microstates A and B, could provide an objective index of negative symptom burden. While clinical translation will require validation in larger cohorts, an objective neurophysiological marker obtainable early in the disease course could be valuable for risk stratification, and for further guiding early targeted intervention strategies (75,76). Classification accuracy may also be improved by combining microstate features with additional neurophysiological features reported to differ in schizophrenia, such as neural responses to mismatch negativity tasks. More broadly, microstate measures may serve as valuable physiological endpoints in intervention studies such as neuromodulation or medication trials (77,78), to assess whether treatment can help to normalise altered neural activity patterns, and whether such changes track symptom improvement.

### Limitations and Future Directions

Several limitations should be considered. First, although the sample size is comparable to many FESSP and schizophrenia microstate studies, it remains modest for multivariate classification and symptom correlations, potentially limiting sensitivity to detect small but clinically meaningful associations. Second, some FESSP participants were taking antipsychotic medications, which may have influenced EEG recordings. However, focusing on FESSP reduces chronicity-related effects and remains informative for probing early pathophysiology. Third, our microstate model was restricted to four canonical classes (A-D), which simplifies interpretability and facilitates comparison with the most commonly reported microstate framework (16,24), and reduces feature dimensionality and multiple-comparisons. This choice may nonetheless oversimplify microstate organisation as some studies have reported additional microstate classes beyond the canonical four (79).

Replication in larger samples with explicit modelling of medication effects and evaluation of alternative microstate model orders will be important considerations for future studies to test the robustness and generalisability of the present results.

### Conclusion

Resting-state EEG microstate dynamics showed a reproducible, multivariate signature that modestly but significantly discriminated the FESSP from HC groups, despite no single feature surviving multiple comparison correction. This pattern suggests that clinically relevant information is distributed across correlated temporal and transition parameters, supporting the use of multivariate decoding approaches to capture modest but complementary effects that are not apparent in univariate tests. Additional exploratory brain-symptom analyses further indicated that negative symptom severity was most closely related to altered microstate organisation, with higher SANS scores associated with reduced temporal stability of microstate D and increased occurrence of microstates A and B. This finding is consistent with models emphasising disrupted top-down coordination and a shift in network state engagement in early psychosis. Overall, these findings support microstate-derived features as scalable electrophysiological markers of early psychosis, while underscoring the need for replication in larger cohorts and more comprehensive designs to establish generalisability, medication sensitivity, and clinical utility.

## Data Availability

This study analysed a pre-collected open dataset from OpenNeuro (Dataset: ds003944).

## Notes

### Competing Interest Statement

The authors have declared no competing interest.

### Funding Statement

This study did not receive any funding.

### Author Declarations

Ethical approval for the original data collection was provided by the University of Pittsburgh International Review Board. Deakin University granted an ethics exemption for secondary analysis.

