## Supplementary Material for "Multivariate Classification of First-Episode Schizophrenia Spectrum Psychosis using EEG Microstate Dynamics"

**Table S1:** Comparison of microstate features (28 features) between the First-Episode Schizophrenia Spectrum Psychosis (FESSP) and Healthy Control (HC) groups

| **feature** | **mean_psychosis** | **mean_control** | **diff_psy_minus_ctrl** | **hedges_g** | **p_ttest** | **p_fdr** |
| --- | --- | --- | --- | --- | --- | --- |
| **MeanDuration_B** | 0.039 | 0.043 | -0.003 | -0.564 | 0.025 | 0.288 |
| **MeanOccurrence_A** | 6.048 | 5.421 | 0.627 | 0.549 | 0.028 | 0.288 |
| **MeanDuration_C** | 0.044 | 0.049 | -0.004 | -0.536 | 0.031 | 0.288 |
| **MeanDuration_A** | 0.041 | 0.045 | -0.004 | -0.528 | 0.057 | 0.338 |
| **MeanOccurrence_C** | 6.441 | 5.888 | 0.553 | 0.474 | 0.06 | 0.338 |
| **MeanOccurrence_D** | 6.08 | 5.576 | 0.504 | 0.417 | 0.075 | 0.348 |
| **MeanDuration_D** | 0.04 | 0.044 | -0.004 | -0.385 | 0.142 | 0.569 |
| **MeanOccurrence_B** | 5.946 | 5.514 | 0.432 | 0.329 | 0.199 | 0.697 |
| **OrgTM_A->C** | 8.883 | 8.539 | 0.344 | 0.151 | 0.523 | 0.993 |
| **OrgTM_C->A** | 8.784 | 8.456 | 0.328 | 0.144 | 0.543 | 0.993 |
| **OrgTM_C->B** | 8.292 | 8.516 | -0.224 | -0.124 | 0.625 | 0.993 |
| **OrgTM_A->B** | 7.955 | 7.76 | 0.195 | 0.116 | 0.643 | 0.993 |
| **IndExpVar_B** | 0.137 | 0.132 | 0.005 | 0.107 | 0.663 | 0.993 |
| **OrgTM_B->C** | 8.326 | 8.493 | -0.167 | -0.094 | 0.708 | 0.993 |
| **OrgTM_D->B** | 7.88 | 8.056 | -0.176 | -0.09 | 0.709 | 0.993 |
| **OrgTM_A->D** | 7.805 | 7.932 | -0.127 | -0.08 | 0.748 | 0.993 |
| **Coverage_A** | 24.628 | 24.32 | 0.308 | 0.058 | 0.823 | 0.993 |
| **OrgTM_D->C** | 9.139 | 9.262 | -0.123 | -0.051 | 0.834 | 0.993 |
| **OrgTM_D->A** | 7.88 | 7.806 | 0.074 | 0.046 | 0.853 | 0.993 |
| **IndExpVar_A** | 0.141 | 0.139 | 0.002 | 0.043 | 0.87 | 0.993 |
| **IndExpVar_D** | 0.184 | 0.187 | -0.003 | -0.04 | 0.877 | 0.993 |
| **IndExpVar_C** | 0.249 | 0.246 | 0.003 | 0.035 | 0.882 | 0.993 |
| **OrgTM_B->D** | 7.828 | 7.894 | -0.066 | -0.033 | 0.889 | 0.993 |
| **Coverage_D** | 24.186 | 24.387 | -0.201 | -0.028 | 0.909 | 0.993 |
| **Coverage_B** | 23.21 | 23.306 | -0.096 | -0.021 | 0.932 | 0.993 |
| **OrgTM_C->D** | 9.26 | 9.311 | -0.051 | -0.021 | 0.932 | 0.993 |
| **OrgTM_B->A** | 7.968 | 7.973 | -0.005 | -0.003 | 0.99 | 0.993 |
| **Coverage_C** | 27.976 | 27.988 | -0.011 | -0.002 | 0.993 | 0.993 |

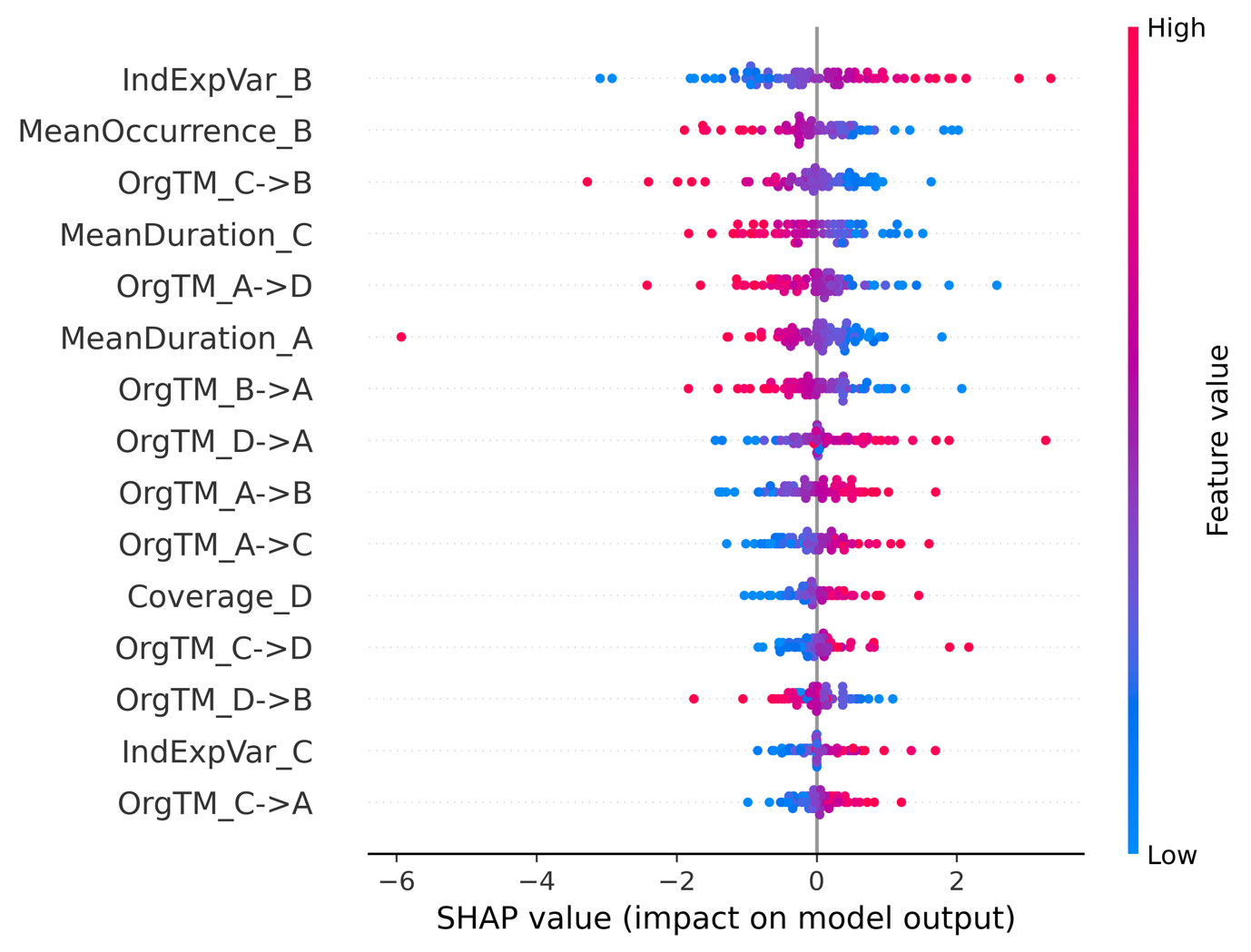

**Figure S1:** SHapley Additive exPlanations (SHAP) beeswarm plot for the linear SVM classifier discriminating first-episode schizophrenia spectrum psychosis (FESSP) from healthy controls (HC). Each point represents one participant. The x-axis shows the SHAP value for a given feature, indicating the direction and magnitude of that feature’s contribution to the SVM decision function for that participant. Positive SHAP values increase the decision function and therefore push the prediction towards the FESSP class, whereas negative SHAP values decrease the decision function and push the prediction towards the HC class. Features are ordered by mean absolute SHAP value across participants (top 15 shown). Point colour indicates the raw feature value (blue=low, red=high). SHAP values were computed out-of-fold within the stratified 5-fold cross-validation procedure.

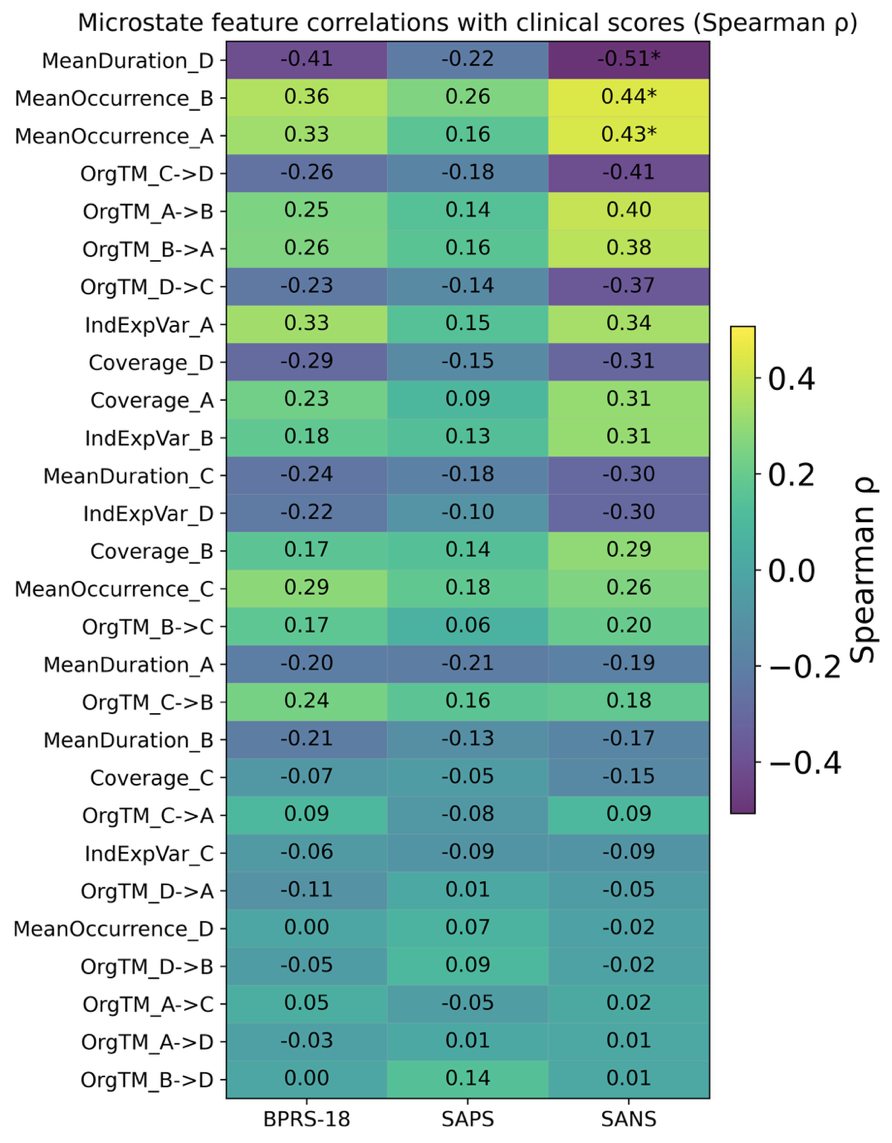

**Figure S2.** Heatmap of microstate feature associations with clinical symptom severity in the First-Episode Schizophrenia Spectrum Psychosis (FESSP) group (N=41). Cell values show Spearman rank correlation coefficients (ρ) between each microstate temporal or transition feature (rows) and symptom scores (columns: BPRS, SAPS, SANS). Features are ordered by the absolute magnitude of the SANS correlation. Asterisks denote associations surviving Benjamini-Hochberg FDR correction across the 28 feature-wise correlations within each clinical scale (pFDR<0.05). BPRS: Brief Psychiatric Rating Scale, SAPS: Scale for the Assessment of Positive Symptoms, SANS: Scale for the Assessment of Negative Symptoms.

**Table S3:** Correlations between microstate features and Brief Psychiatric Rating Scale (BPRS) scores for the First-Episode Schizophrenia Spectrum Psychosis (FESSP) group.

| **Feature** | **N** | **Spearman ρ** | **p** | **p (FDR-BH)** |
| --- | --- | --- | --- | --- |
| **MeanDuration_D** | 41 | -0.41 | 0.008 | 0.215 |
| **MeanOccurrence_B** | 41 | 0.364 | 0.019 | 0.257 |
| **IndExpVar_A** | 41 | 0.329 | 0.036 | 0.257 |
| **MeanOccurrence_A** | 41 | 0.327 | 0.037 | 0.257 |
| **Coverage_D** | 41 | -0.293 | 0.063 | 0.319 |
| **MeanOccurrence_C** | 41 | 0.286 | 0.07 | 0.319 |
| **OrgTM_B->A** | 41 | 0.26 | 0.101 | 0.319 |
| **OrgTM_C->D** | 41 | -0.258 | 0.103 | 0.319 |
| **OrgTM_A->B** | 41 | 0.252 | 0.113 | 0.319 |
| **MeanDuration_C** | 41 | -0.243 | 0.126 | 0.319 |
| **OrgTM_C->B** | 41 | 0.24 | 0.131 | 0.319 |
| **OrgTM_D->C** | 41 | -0.229 | 0.15 | 0.319 |
| **Coverage_A** | 41 | 0.227 | 0.153 | 0.319 |
| **IndExpVar_D** | 41 | -0.224 | 0.159 | 0.319 |
| **MeanDuration_B** | 41 | -0.212 | 0.183 | 0.341 |
| **MeanDuration_A** | 41 | -0.203 | 0.203 | 0.354 |
| **IndExpVar_B** | 41 | 0.182 | 0.256 | 0.421 |
| **OrgTM_B->C** | 41 | 0.173 | 0.28 | 0.436 |
| **Coverage_B** | 41 | 0.166 | 0.299 | 0.441 |
| **OrgTM_D->A** | 41 | -0.106 | 0.51 | 0.713 |
| **OrgTM_C->A** | 41 | 0.092 | 0.568 | 0.758 |
| **Coverage_C** | 41 | -0.072 | 0.656 | 0.834 |
| **IndExpVar_C** | 41 | -0.063 | 0.695 | 0.846 |
| **OrgTM_D->B** | 41 | -0.048 | 0.763 | 0.861 |
| **OrgTM_A->C** | 41 | 0.047 | 0.769 | 0.861 |
| **OrgTM_A->D** | 41 | -0.033 | 0.838 | 0.903 |
| **MeanOccurrence_D** | 41 | 0.003 | 0.987 | 0.989 |
| **OrgTM_B->D** | 41 | 0.002 | 0.989 | 0.989 |

**Table S4:** Correlations between microstate features and Scale for the Assessment of Positive Symptoms (SAPS) scores for the First-Episode Schizophrenia Spectrum Psychosis (FESSP) group.

| **Feature** | **N** | **Spearman ρ** | **p** | **p (FDR-BH)** |
| --- | --- | --- | --- | --- |
| **MeanOccurrence_B** | 41 | 0.258 | 0.103 | 0.712 |
| **MeanDuration_D** | 41 | -0.221 | 0.165 | 0.712 |
| **MeanDuration_A** | 41 | -0.214 | 0.18 | 0.712 |
| **MeanDuration_C** | 41 | -0.183 | 0.252 | 0.712 |
| **MeanOccurrence_C** | 41 | 0.18 | 0.261 | 0.712 |
| **OrgTM_C->D** | 41 | -0.176 | 0.272 | 0.712 |
| **OrgTM_C->B** | 41 | 0.165 | 0.303 | 0.712 |
| **MeanOccurrence_A** | 41 | 0.162 | 0.312 | 0.712 |
| **OrgTM_B->A** | 41 | 0.157 | 0.327 | 0.712 |
| **IndExpVar_A** | 41 | 0.146 | 0.363 | 0.712 |
| **Coverage_D** | 41 | -0.145 | 0.365 | 0.712 |
| **OrgTM_A->B** | 41 | 0.144 | 0.37 | 0.712 |
| **OrgTM_D->C** | 41 | -0.143 | 0.372 | 0.712 |
| **Coverage_B** | 41 | 0.142 | 0.377 | 0.712 |
| **OrgTM_B->D** | 41 | 0.139 | 0.386 | 0.712 |
| **IndExpVar_B** | 41 | 0.127 | 0.429 | 0.712 |
| **MeanDuration_B** | 41 | -0.126 | 0.432 | 0.712 |
| **IndExpVar_D** | 41 | -0.098 | 0.544 | 0.764 |
| **OrgTM_D->B** | 41 | 0.093 | 0.561 | 0.764 |
| **IndExpVar_C** | 41 | -0.093 | 0.563 | 0.764 |
| **Coverage_A** | 41 | 0.091 | 0.573 | 0.764 |
| **OrgTM_C->A** | 41 | -0.079 | 0.625 | 0.795 |
| **MeanOccurrence_D** | 41 | 0.071 | 0.658 | 0.801 |
| **OrgTM_B->C** | 41 | 0.062 | 0.699 | 0.816 |
| **Coverage_C** | 41 | -0.049 | 0.763 | 0.828 |
| **OrgTM_A->C** | 41 | -0.047 | 0.769 | 0.828 |
| **OrgTM_D->A** | 41 | 0.015 | 0.928 | 0.947 |
| **OrgTM_A->D** | 41 | 0.011 | 0.947 | 0.947 |

**Table S5:** Correlations between microstate features and Scale for the Assessment of Negative Symptoms (SANS) scores for the First-Episode Schizophrenia Spectrum Psychosis (FESSP) group.

| **Feature** | **N** | **Spearman ρ** | **p** | **p (FDR-BH)** |
| --- | --- | --- | --- | --- |
| **MeanDuration_D** | 41 | -0.507 | 0.001 | 0.02 |
| **MeanOccurrence_B** | 41 | 0.443 | 0.004 | 0.042 |
| **MeanOccurrence_A** | 41 | 0.434 | 0.005 | 0.042 |
| **OrgTM_C->D** | 41 | -0.41 | 0.008 | 0.054 |
| **OrgTM_A->B** | 41 | 0.399 | 0.01 | 0.055 |
| **OrgTM_B->A** | 41 | 0.38 | 0.014 | 0.067 |
| **OrgTM_D->C** | 41 | -0.366 | 0.019 | 0.075 |
| **IndExpVar_A** | 41 | 0.341 | 0.029 | 0.103 |
| **Coverage_D** | 41 | -0.311 | 0.048 | 0.12 |
| **Coverage_A** | 41 | 0.308 | 0.05 | 0.12 |
| **IndExpVar_B** | 41 | 0.307 | 0.051 | 0.12 |
| **MeanDuration_C** | 41 | -0.302 | 0.055 | 0.12 |
| **IndExpVar_D** | 41 | -0.301 | 0.056 | 0.12 |
| **Coverage_B** | 41 | 0.293 | 0.063 | 0.125 |
| **MeanOccurrence_C** | 41 | 0.26 | 0.101 | 0.188 |
| **OrgTM_B->C** | 41 | 0.198 | 0.214 | 0.375 |
| **MeanDuration_A** | 41 | -0.187 | 0.241 | 0.386 |
| **OrgTM_C->B** | 41 | 0.185 | 0.248 | 0.386 |
| **MeanDuration_B** | 41 | -0.168 | 0.294 | 0.434 |
| **Coverage_C** | 41 | -0.147 | 0.359 | 0.503 |
| **OrgTM_C->A** | 41 | 0.091 | 0.57 | 0.743 |
| **IndExpVar_C** | 41 | -0.088 | 0.584 | 0.743 |
| **OrgTM_D->A** | 41 | -0.05 | 0.757 | 0.922 |
| **MeanOccurrence_D** | 41 | -0.021 | 0.898 | 0.969 |
| **OrgTM_D->B** | 41 | -0.019 | 0.904 | 0.969 |
| **OrgTM_A->C** | 41 | 0.017 | 0.918 | 0.969 |
| **OrgTM_A->D** | 41 | 0.007 | 0.966 | 0.969 |
| **OrgTM_B->D** | 41 | 0.006 | 0.969 | 0.969 |
